# Examining ChatGPT’s Ability to Answer Cirrhosis Related Questions in Arabic Compared to English

**DOI:** 10.1101/2023.07.05.23292147

**Authors:** Jamil S. Samaan, Yee Hui Yeo, Wee Han Ng, Peng-Sheng Ting, Hirsh Trivedi, Aarshi Vipani, Ju Dong Yang, Omer Liran, Brennan Spiegel, Alexander Kuo, Walid Ayoub

## Abstract

**Background and Study Aims:** Cirrhosis is a chronic progressive disease which requires complex care. Its incidence is rising in the Arab countries making it the 7^th^ leading cause of death in the Arab League in 2010. ChatGPT is a large language model with a growing body of literature demonstrating its ability to answer clinical questions. We examined ChatGPT’s accuracy in responding to cirrhosis related questions in Arabic and compared its performance to English.

**Materials and Methods:** ChatGPTs responses to 91 questions in Arabic and English were graded by a transplant hepatologist fluent in both languages. Accuracy of responses was assessed using the scale: 1. Comprehensive, 2. Correct but inadequate, 3. Mixed with correct and incorrect/outdated data, and 4. Completely incorrect. Accuracy of Arabic compared to English responses was assessed using the scale: 1. Arabic response is more accurate, 2. Similar accuracy, 3. Arabic response is less accurate.

**Results:** The model provided 22 (24.2%) comprehensive, 44 (48.4%) correct but inadequate, 13 (14.3%) mixed with correct and incorrect/outdated data and 12 (13.2%) completely incorrect Arabic responses. When comparing the accuracy of Arabic and English responses, 9 (9.9%) of the Arabic responses were graded as more accurate, 52 (57.1%) similar in accuracy and 30 (33.0%) as less accurate compared to English.

**Conclusion:** ChatGPT has the potential to serve as an adjunct source of information for Arabic speaking patients with cirrhosis. The model provided correct responses in Arabic to 72.5% of questions, although its performance in Arabic was less accurate than in English. The model produced completely incorrect responses to 13.2% of questions, reinforcing its potential role as an adjunct and not replacement of care by licensed healthcare professionals. Future studies to refine this technology are needed to help Arabic speaking patients with cirrhosis across the globe understand their disease and improve their outcomes.

## INTRODUCTION

Chat Generative Pre-trained Transformer (ChatGPT) is a large language model which was released by the company OpenAI in November 2022 and has exponentially grown in popularity among the general public worldwide^1^. This model was trained on a large dataset from a broad range of topics although the exact data source is unknown. It has the capability to comprehend user questions and respond with easy to understand, conversational and seemingly knowledgeable answers. A recent study examined the knowledge of ChatGPT related to Cirrhosis and Hepatocellular Carcinoma and found the model answered 79% of questions with correct responses, 47% of which were graded as comprehensive^2^. Its ability to answer clinical questions has also been examined in other topics such as bariatric surgery and cancer^3,4^. These studies utilized the English language when prompting ChatGPT, while data regarding its ability to comprehend and respond in other languages is limited.

Cirrhosis is a chronic progressive disease where adherence to medication regimens, lifestyle modifications, regular follow up and routine cancer screening is critical to achieving optimal outcomes. Given the complexity of care required to manage this disease, patient health knowledge is essential, with one study showing an association between poor health literacy and poor liver function^5^. Furthermore, the usefulness of information from online health platforms and large hepatology centers in the United States, Europe and Asia may be limited by lengthy text and complexity of language^6^. Although most of the large hepatology centers publish information in English, a large portion of patients seeking such information don’t speak or understand English. Considering these challenges, large language models such as ChatGPT may serve as a valuable easy to understand, quick and accessible source of information for patients with cirrhosis.

Cirrhosis is on the rise in the Arab countries and ranked the 7^th^ leading cause of death in the Arab League during the year 2010^7^. Furthermore, displacement and migration of citizens from the middle east sharply rose from 2005 to 2015 with doubling of migrant populations from the region^8^. Large language models may be leveraged to help citizens of Arab countries as well as bridge language barriers for Arabic speaking migrants around the world who have cirrhosis. We examined the ability of ChatGPT to understand liver cirrhosis related questions written in Arabic and assess the accuracy of its responses. To put its performance in context, we compared ChatGPT’s performance when responding to English and Arabic questions.

## MATERIALS AND METHODS

### Question Curation

A total of 91 questions were included in our study. Patient questions related to liver cirrhosis were collected from professional societies and institutions. Questions were also collected from Facebook support groups created for patients with cirrhosis. Details regarding question curation and selection are described elsewhere^2^. Questions were then translated into Arabic and verified by two authors who are fluent in Arabic and English (JS, WA).

### Response Generation and Grading

ChatGPT is a large language model which was trained on a large dataset containing a broad range of topics. The model was trained using a technique called Reinforcement Learning from Human Feedback or Reinforcement Learning from Human Preference (RLHF/RLHP). This allows the model to fine tube its responses in a way that is coherent and conversational by using human feedback^9^. Each question was entered into the ChatGPT January 30^th^ version in both Arabic and English and responses were recorded. Each question was entered as an individual prompt using the “New Chat” function. The Arabic responses were then graded for accuracy by a board-certified transplant hepatologist reviewer who is fluent in English and Arabic and has more than 15 years of clinical experience in transplant hepatology (WA). The following grading scale was used to grade each response: 1. Comprehensive, 2. Correct but inadequate, 3. Mixed with correct and incorrect/outdated data, and 4. Completely incorrect. The same reviewer then assessed the quality of the Arabic compared to English responses by using the following scale: 1. Arabic response is more accurate, 2. Similar accuracy, 3. Arabic response is less accurate.

### Statistical analysis

For statistical analysis purposes, questions were categorized into multiple subgroups: Basic knowledge, diagnosis, treatment, lifestyle, preventative care and others. Proportion of grades were presented as percentages overall as well as stratified by subgroups. Microsoft Excel (version 16.69.1) was used for all analysis.

## RESULTS

A total of 91 questions were included in our study. When examining the accuracy of Arabic responses, the model provided 22 (24.2%) comprehensive, 44 (48.4%) correct but inadequate, 13 (14.3%) mixed with correct and incorrect/outdated data and 12 (13.2%) completely incorrect responses. When stratified by subgroups, the model performed best in the lifestyle subgroup with 36.4% of responses graded as Comprehensive and 45.5% graded as correct but inadequate (Table 1). The diagnosis and treatment subgroups contained the highest proportion of completely incorrect responses with 33.3% and 31.3%, respectively, although the sample size was small in the diagnosis subgroup (n=3).

**Table 1.** Grading of responses generated by ChatGPT to Arabic questions related to cirrhosis categorized by subgroup.

|  |  |
| --- | --- |
| **Basic Knowledge (N=36)** |  |
| 1. Comprehensive | 19.4% |
| 2. Correct but inadequate | 55.6% |
| 3. Mixed with correct and incorrect/outdated data | 8.3% |
| 4. Completely incorrect | 16.7% |
| **Diagnosis (N=3)** |  |
| 1. Comprehensive | 0% |
| 2. Correct but inadequate | 66.7% |
| 3. Mixed with correct and incorrect/outdated data | 0% |
| 4. Completely incorrect | 33.3% |
| **Treatment (N=16)** |  |
| 1. Comprehensive | 12.5% |
| 2. Correct but inadequate | 31.3% |
| 3. Mixed with correct and incorrect/outdated data | 25.0% |
| 4. Completely incorrect | 31.3% |
| **Lifestyle (N=22)** |  |
| 1. Comprehensive | 36.4% |
| 2. Correct but inadequate | 45.5% |
| 3. Mixed with correct and incorrect/outdated data | 18.2% |
| 4. Completely incorrect | 0% |
| **Preventative Care (N=4)** |  |
| 1. Comprehensive | 0% |
| 2. Correct but inadequate | 50.0% |
| 3. Mixed with correct and incorrect/outdated data | 50.0% |
| 4. Completely incorrect | 0% |
| **Others (N=10)** |  |
| 1. Comprehensive | 50.0% |
| 2. Correct but inadequate | 50.0% |
| 3. Mixed with correct and incorrect/outdated data | 0% |
| 4. Completely incorrect | 0% |

When comparing the accuracy of Arabic and English responses, 9 (9.9%) of the Arabic responses were graded as more accurate, 52 (57.1%) similar in accuracy and 30 (33.0%) as less accurate compared to English. The model performed best in Arabic compared to English in the preventative care subgroup with 25.0% of responses in Arabic graded as more accurate than English (Table 2). The diagnosis and basic knowledge subgroups contained the highest rate of the grade “Arabic is less accurate than English” with 66.7% and 44.4%, respectively, although the sample size was small in the diagnosis subgroup (n=3). For example the Arabic and English responses provided similar answers on the approach to the management of cirrhosis and vaccination. The Arabic version, however, provided a better explanation for “what is cirrhosis”. Unlike the English version, the Arabic version failed to give an accurate description of varices related to cirrhosis.

**Table 2.** Grading of responses comparing the accuracy between Arabic and English responses generated by ChatGPT to Cirrhosis related questions categorized by subgroup.

|  |  |
| --- | --- |
| **Basic Knowledge (N=36)** |  |
| 1. Arabic response is more accurate | 2.8% |
| 2. Similar accuracy | 52.8% |
| 3. Arabic response is less accurate | 44.4% |
| **Diagnosis (N=3)** |  |
| 1. Arabic response is more accurate | 0% |
| 2. Similar accuracy | 33.3% |
| 3. Arabic response is less accurate | 66.7% |
| **Treatment (N=16)** |  |
| 1. Arabic response is more accurate | 12.5% |
| 2. Similar accuracy | 43.8% |
| 3. Arabic response is less accurate | 43.8% |
| **Lifestyle (N=22)** |  |
| 1. Arabic response is more accurate | 13.6% |
| 2. Similar accuracy | 72.7% |
| 3. Arabic response is less accurate | 13.6% |
| **Preventative Care (N=4)** |  |
| 1. Arabic response is more accurate | 25.0% |
| 2. Similar accuracy | 50.0% |
| 3. Arabic response is less accurate | 25.0% |
| **Others (N=10)** |  |
| 1. Arabic response is more accurate | 20.0% |
| 2. Similar accuracy | 70.0% |
| 3. Arabic response is less accurate | 10.0% |

## DISCUSSION

The large language model ChatGPT has made patient access to artificial intelligence easier than ever and mainstream. Its ability to comprehend clinical questions and provide easy to understand and seemingly knowledgeable answers will likely make it an attractive source for patients seeking information related to their medical care. Our study builds on the current literature showing the potential utility of ChatGPT as an adjunct source of information for patients by describing its ability to answer questions related to cirrhosis in Arabic. The model’s performance in Arabic was impressive, providing correct responses to 72.5% of questions, 24.2% of which were graded as “comprehensive”. When comparing the accuracy of Arabic responses to English, Arabic responses were similar 57.1% the time but less accurate 33.0% of the time, highlighting a disparity in performance. Given the rapid advancement of this technology since its release in November of 2022, it has the potential to serve as a valuable adjunct source of information for Arabic speaking patients around the world.

Language barriers in medicine have been previously described with studies showing worse outcomes for patients that have language discordance with their healthcare provider ^10,11^. Furthermore, patient’s use of online resources for medical information is on the rise with one in three individuals in the United States having searched online to figure out a medical condition and one half of European Union citizens reported using the internet for information related to injury, disease, nutrition, improving health or similar ^12,13^. Navigating search engines can be a daunting task for patients, from the time needed to find information, to identifying reliable sources and misinformation. We anticipate the use of large language models such as ChatGPT will significantly increase by patients due to its ease of use and quick access interface as well as its simple and conversational responses. This provides great urgency across the medical field to examine the utility but more importantly the limitations of this technology in order to better counsel patients. Our results are promising given the impressive performance of ChatGPT in Arabic. We anticipate significant improvement in future versions of ChatGPT and with the development of larger and more powerful language models given the rapid growth and advancement in this field.

ChatGPT has important limitations to consider. One important concept is the “hallucinations” or stochastic phenomena where the model produces incorrect statements while sounding confident, leading to increased risk of spreading misinformations^14^. This is demonstrated in our study by the model providing “completely incorrect” answers to 14.3% of questions despite sounding confident and knowledgeable. Due to this important limitation, we emphasize the role of ChatGPT as a potential adjunct and not replacement to care provided by licensed healthcare professionals. There also appears to be a difference in performance in Arabic compared to English, as evident by the 33.0% of responses in Arabic graded as less accurate than English. The cause of this is unclear although we hypothesize that the source of information utilized by the model may be an important factor and is unknown at the current time. Generally there is more English literature related to diagnosis and treatment compared to lifestyle and basic knowledge which may have contributed to the disparity. It is also unclear if the model is using sources in Arabic to answer questions in Arabic, or translating information from its dataset into Arabic. Future studies examining differences in performance in languages other than English may help shed light on this disparity.

## CONCLUSION

ChatGPT has the potential to serve as an adjunct source of information for Arabic speaking patients with cirrhosis. The model provided correct responses in Arabic to 72.5% of questions, 24.2% of which were graded as comprehensive, although its performance in Arabic was less accurate than in English. While its performance was impressive, the model did produce completely incorrect responses to 13.2% of questions, reinforcing its potential role as an adjunct and not replacement to care by licensed healthcare professionals. Future studies to refine this technology are needed to help Arabic speaking patients with cirrhosis across the globe understand their disease and improve their outcomes.

## Data Availability

All data produced in the present study are available upon reasonable request to the authors.

## Acknowledgements

None.

## Notes

**Conflict of Interest Statement:** None declared.

**Funding/Support:** This research did not receive any specific grant from funding agencies in the public, commercial or not-for-profit sectors.

### Competing Interest Statement

The authors have declared no competing interest.

### Funding Statement

This study did not receive any funding

